# Quantifying the Impact of Ocrelizumab on Paramagnetic Rim Lesions in Multiple Sclerosis

**DOI:** 10.1101/2025.05.20.25328011

**Authors:** Kimberly H. Markowitz, Neha V. Safi, Alexey Dimov, Ulrike W. Kaunzner, Ha Luu, Mert Sisman, Yi Wang, Thanh D. Nguyen, Sandra Hurtado Rúa, Susan A. Gauthier

## Abstract

Paramagnetic rim lesions (PRLs) are a subset of chronic active multiple sclerosis (MS) lesions, characterized by a rim of iron-laden macrophages and microglia encircling a demyelinated core. These lesions signify persistent inflammatory activity in the central nervous system (CNS), yet their responsiveness to B cell depletion therapy has not been well established. Ocrelizumab, a humanized monoclonal antibody that targets CD20+ B cells, is effective in suppressing acute MS activity; however, its impact on chronic CNS inflammation, particularly within PRLs, remains unclear. To address this question, we conducted a retrospective longitudinal imaging study of MS patients undergoing Ocrelizumab treatment. All participants had pre-treatment imaging with at least one PRL identified via quantitative susceptibility mapping (QSM). Using an R2*QSM source separation (SS) framework, we disentangled the contribution from iron sources with positive susceptibility (QSMp) and myelin sources with negative susceptibility (QSMn) to quantify iron and myelin changes within lesions. Lesions were individually segmented, and PRLs were classified through expert consensus. Modeling of QSMp and QSMn was performed using mixed effects and joint-point regression analysis to assess temporal changes in lesion properties. Across 29 patients, we identified 97 PRLs. Prior to treatment, PRLs exhibited significantly elevated QSMp compared to non-PRLs (p=0.001), consistent with iron enrichment at the lesion rim. Following Ocrelizumab initiation, PRLs showed a significantly greater reduction in QSMp than non-PRLs (p<0.001), with joint-point analysis revealing an accelerated decline beginning approximately four months after treatment for PRLs. PRLs demonstrated lower QSMn values than non-PRLs pre-treatment (p<0.001), but without significant post-treatment divergence (p=0.382). Joint-point analysis revealed a steep increase in QSMn beginning approximately three months after treatment for PRLs. These findings suggest that Ocrelizumab may influence the trajectory of chronic active MS lesions by attenuating iron-related inflammation. This work provides preliminary insight into the potential of B cell-targeting therapies to modulate slow-burning inflammatory processes in MS and underscores the value of advanced MRI techniques, such as R2*QSM SS, in capturing subtle treatment effects on chronic CNS pathology.

## Introduction

Multiple Sclerosis (MS) is a chronic autoimmune disease of the central nervous system (CNS) characterized by white and gray matter demyelination and pro-inflammatory immune activity.^1^ Active demyelinating lesions, driven by blood-brain barrier (BBB) disruption and infiltration of T and B lymphocytes into the CNS, are detectable with gadolinium-enhanced T1-weighted imaging.^2–4^ However, as the BBB restores, chronic active lesions with ongoing demyelination at the lesion edge^5^ become undetectable using gadolinium-enhanced imaging, necessitating alternative imaging techniques.^6^ Among these chronic active lesions, paramagnetic rim lesions (PRLs) are particularly clinically significant. Defined by a demyelinated core surrounded by iron-laden macrophages and microglia, PRLs reflect persistent inflammation and tissue damage at the lesion rim.^4,7^ PRLs are associated with a more aggressive disease course, increased disability, and faster disease progression,^8–10^ emphasizing the need for effective therapeutic interventions targeting these lesions^11^ and the underlying mechanisms driving their formation.

One of the key drivers of MS pathology is the role of B cells in promoting CNS inflammation and damage through multiple mechanisms, including antigen presentation, pro-inflammatory cytokine release, and antibody production.^12^ Therapeutic approaches targeting B cells, such as Ocrelizumab, have demonstrated substantial efficacy in modifying acute disease activity.^13^ Ocrelizumab, a monoclonal antibody approved for both relapsing-remitting MS and primary progressive MS, selectively depletes CD20+ B cells in the periphery by binding to the CD20 glycoprotein and is suggested to induce apoptosis through cellular cytotoxicity.^14^ However, its potential secondary effect on chronic inflammation within PRLs, particularly behind a restored BBB, remains unclear.^15^

Quantitative susceptibility mapping (QSM), a technique that extracts tissue magnetic susceptibility by deconvolving the phase data from the gradient echo imaging (GRE) has been used to identify and quantify the underlying pathology of PRLs.^6,16,17^ While QSM signals reflect contributions from both paramagnetic iron and diamagnetic myelin, source separation (SS) algorithms can differentiate between these components within a voxel, offering detailed insights into these concurrent processes. The R2*QSM framework leverages the proportionality between susceptibility and the R2* decay rate to enable separation of iron sources with positive susceptibility (QSMp) and myelin sources with negative susceptibility (QSMn) that colocalize within an imaging voxel. This algorithm provides perfectly aligned iron and myelin maps from a single GRE data acquisition, offering a significant scanning advantage over multiple acquisition methods.^18,19^

In this study, we aimed to determine whether treatment using Ocrelizumab has a potential downstream effect on the CNS by altering the trajectory of chronic inflammation in PRLs. To do this, we conducted a longitudinal analysis of susceptibility change across both iron and myelin components, utilizing the R2*QSM SS framework.

## Materials and Methods

### Patient Cohort

This is a retrospective single-arm study designed to evaluate the impact of Ocrelizumab treatment on PRLs using QSM SS. Participants were selected retrospectively from an ongoing clinical and imaging database at the Weill Cornell MS Center. Inclusion criteria for this study were: (1) the diagnosis of clinically isolated syndrome (CIS) or MS meeting the 2010 revised McDonald criteria^20^; (2) subjects currently participating in the clinical and MRI MS research repository and ≥ 18 years old; (3) started Ocrelizumab and had at least one year of prior imaging; (4) had at least one PRL defined on QSM. Demographic and clinical data including age, sex, disease subtype, disease duration from date of first symptom, previous disease modifying treatment (DMT), and expanded disability status scale (EDSS) were collected. DMTs were divided into lower and higher efficacy groups. Lower efficacy DMTs included: interferon beta-1a, glatiramer acetate, and dimethyl fumarate. Higher efficacy DMTs included: fingolimod and natalizumab. Clinical data were collected for all available pre- and post-treatment MRI time points available.

### MRI Acquisition and Reconstruction

Longitudinal imaging was performed at 3T using GE Signa HDxt (GE Healthcare, Waukesha, WI, USA) and Siemens Magnetom Skyra (Siemens Medical Solutions, Malvern, PA, USA) MRI scanners. The MRI protocol consisted of sagittal 3D T1-weighted (T1w) sequence for anatomical structure, 2D T2-weighted (T2w) fast spin echo, and 3D T2w fluid attenuated inversion recovery (FLAIR) sequences for lesion detection, gadolinium-enhanced 3D T1w sequence for acute lesion identification, and axial 3D multi-echo GRE sequence for QSM. Detailed imaging parameters for each scanner are provided below.

*<u>The Siemens scanning protocol consisted of the following sequences:</u>* 1) 3D sagittal T1w MPRAGE: Repetition Time (TR)/Echo Time (TE)/Inversion Time (TI) = 2300/2.3/900 ms, flip angle (FA) = 8°, GRAPPA parallel imaging factor (R) = 2, voxel size = 1.0 × 1.0 × 1.0 mm^3^; 2) 2D axial T2-weighted (T2w) turbo spin echo: TR/TE = 5840/93 ms, FA = 90°, echo train length (ETL) = 18, R = 2, number of signal averages (NSA) = 2, voxel size = 0.5 × 0.5 × 3 mm^3^; 3) 3D sagittal fat-saturated T2w fluid attenuated inversion recovery (FLAIR) SPACE: TR/TE/TI = 8500/391/2500 ms, FA = 90°, ETL = 278, R = 4, voxel size = 1.0 × 1.0 × 1.0 mm^3^.

*<u>The GE scanning protocol consisted of the following sequences:</u>* 1) 3D sagittal T1w BRAVO: TR/TE/TI = 8.8/3.4/450 ms, FA = 15°, voxel size = 1.2 × 1.2 × 1.2 mm^3^, ASSET parallel imaging acceleration factor (R) = 1.5; 2) 2D axial T2w fast spin echo: TR/TE = 5267/86 ms, FA = 90°, ETL = 23, number of excitations (NEX) = 2, voxel size = 0.5 x 0.5 x 3.0 mm^3^; 3) 3D sagittal T2w FLAIR CUBE: TR/TE/TI = 5000/139/1577 ms, FA = 90°, ETL = 162, R = 1.6, voxel size = 1.2 × 1.2 × 1.2 mm^3^.

Both scanners had similar parameters for the *<u>axial 3D multi-echo GRE sequence for QSM</u>:* axial field of view (FOV) = 24 cm, TR/TE1/ΔTE = 48.0/6.3/4.1 ms, number of TEs = 10, FA = 15°, R = 2, voxel size = 0.75 x 0.93 x 3 mm^3^, scan time = 4.2 min. The harmonized QSM imaging protocol has demonstrated high reproducibility across different scanner vendors^21–23^. QSM was reconstructed from complex GRE images using a fully automated Morphology Enabled Dipole Inversion algorithm zero-referenced to the cerebrospinal fluid across the entire brain (MEDI+0)^24^.

For each time point, T1w image was brain-extracted, intensity normalized and aligned to the FreeSurfer T1w conformed space (256 x 256 x 256 1 mm isotropic voxels) using FreeSurfer v6.^25^ Subsequently, T2w, T2w FLAIR, QSM, QSMp and QSMn images acquired at each time point were co-registered to T-1 FreeSurfer (time point immediately preceding treatment initiation) conformed space using ANTs image registration toolbox^26^.

### Reconstruction of Source-Separation Maps

We applied R2*QSM susceptibility source separation algorithm^18,19^ to distinguish susceptibility contribution of positive sources (predominantly iron) from that of negative sources (predominantly myelin) within an imaging voxel. Examples of positive (QSMp) and negative (QSMn) SS maps are shown in Figures 1B and 2C, respectively.

**Figure 1.**
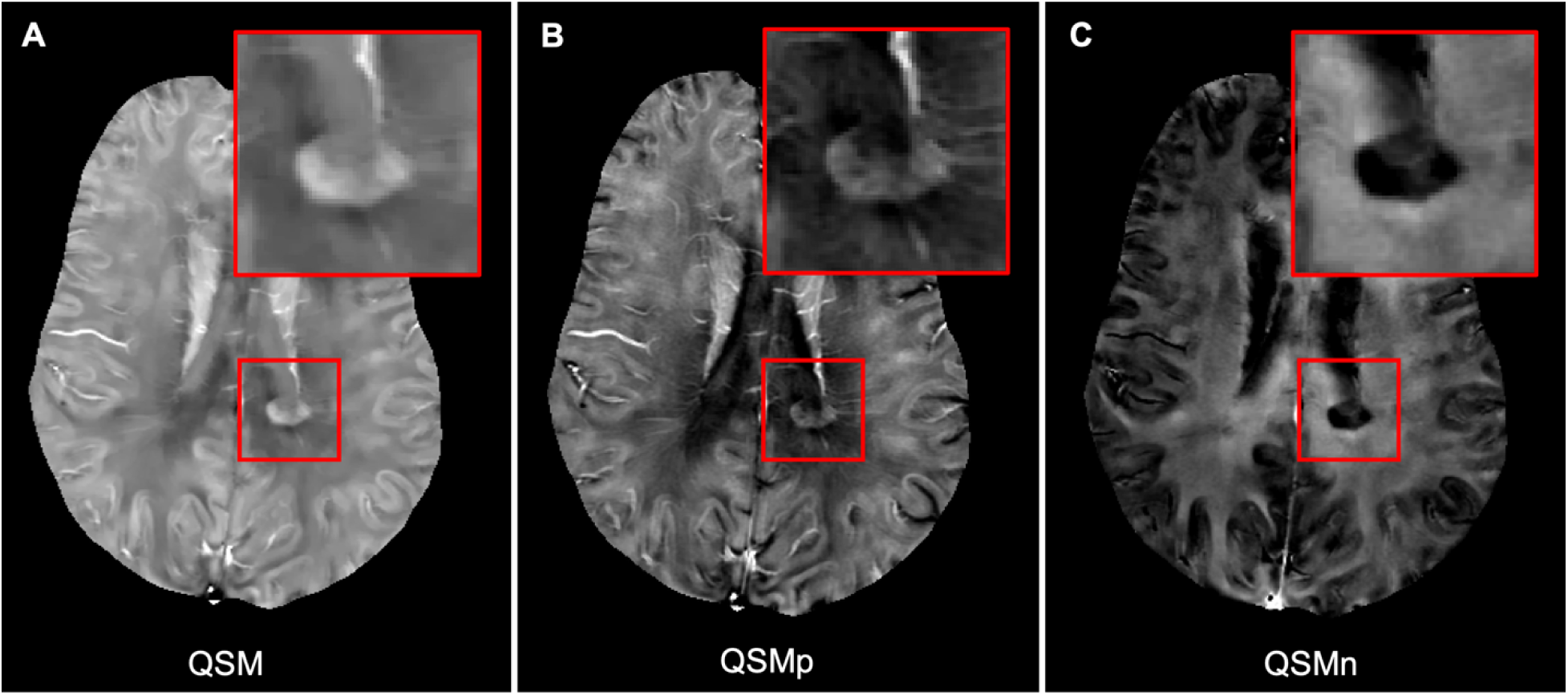
Example of a paramagnetic rim lesion in a patient with RRMS. (A) Quantitative susceptibility mapping (QSM) image reflecting the composite contributions of iron and myelin. (B) QSMp map highlighting paramagnetic material, where greater hyperintensity (higher susceptibility) indicates increased iron deposition. (C) QSMn map highlighting diamagnetic material, where more hypointensity (lower susceptibility) indicates myelin loss.

**Figure 2.**
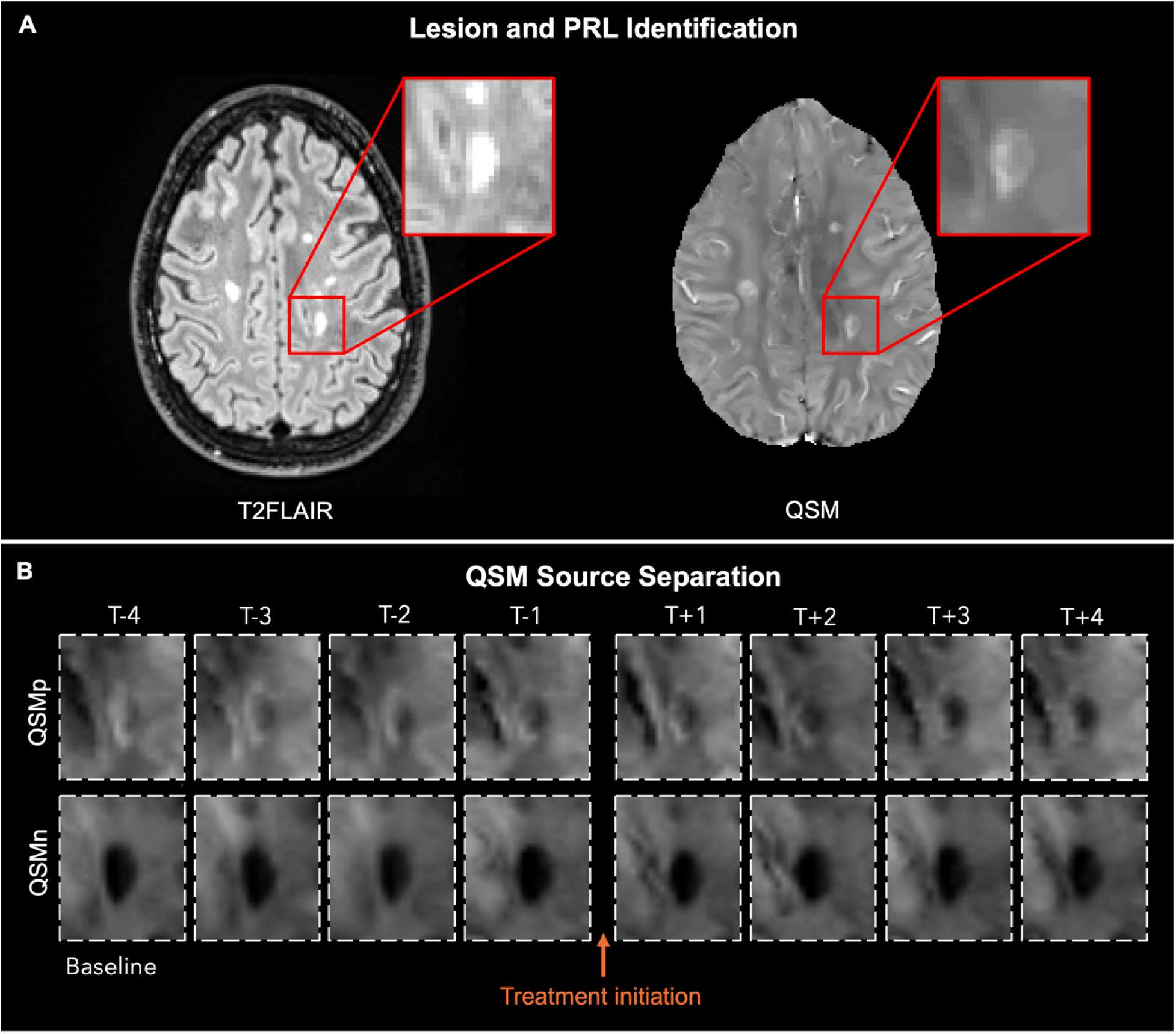
Longitudinal lesion tracking and image postprocessing. (A) Lesion masks were generated and edited based on the baseline T2FLAIR, which corresponds to the patient’s earliest available scan. The T2FLAIR lesion mask, T2FLAIR image, and QSM image were all co-registered to T1 space. The co-registered T2FLAIR lesion label was used to identify PRLs on QSM. The lesion of interest is magnified (inset) to highlight the rim of increased susceptibility. (B) Temporal progression of the lesion was evaluated using QSM and its SS components across all time points (pre- and post-treatment) relative to treatment initiation. All follow-up scans were co-registered to T-1 FreeSurfer (time point immediately preceding treatment initiation), enabling consistent overlay of the baseline lesion mask. QSM and SS data were then extracted from this aligned space for each time point. Top row: total QSM signal (QSM). Middle row: paramagnetic component (QSMp). Bottom row: diamagnetic component (QSMn).

### Lesion Identification and Susceptibility Analysis

MS lesions were automatically identified and segmented on the T2w FLAIR image using the automated Anatomic Information and Lesion-Wise Loss Function (ALL-Net),^27^ which was followed by manual editing and creation of individual lesion labels. T1w and T2w images were referenced, as needed, during the editing process to define lesion boundaries, especially in the case of confluent lesions. The consensus of two blinded reviewers was used to identify PRLs on QSM, and a third independent reviewer resolved any discrepant lesions.^28^ T2w FLAIR lesion labels were co-registered to QSM, QSMp, and QSMn images and edited as needed to ensure accurate depiction of lesion boundary. Acute enhancing lesions on T1w+Gd images were excluded from analysis. Figure 1 provides an example of PRL visualized on QSM, QSMp, and QSMn. The mean QSMp and QSMn values were recorded for each lesion on the baseline and across all co-registered follow-up QSM images (Fig. 2).

### Statistical Analysis

We used a model-based approach to study the trajectory of chronic inflammation in PRLs. We conducted a longitudinal analysis of changes in susceptibility across both iron and myelin components, utilizing the R2*QSM SS framework. The model-based approach includes a mixed effects model and a joint-point regression model.

The mixed effects model^29^ estimated the effect of pre- and post-treatment periods on QSM biomarkers while accounting for multiple lesions per patient. The model was fitted using restricted maximum likelihood (REML), and its fit was assessed through likelihood ratio tests, Akaike information criterion (AIC), and residual diagnostics. Random intercept variance estimates were examined to ensure appropriate within-subject correlation modeling.

A linear joint-point regression model estimated the change points and slopes of the QSM trajectories for PRLs over time. A joint-point model allows detecting structural changes in a continuous response variable by estimating breakpoints where the trajectory shifts.^30^ We implemented both a classical and Bayesian joint-point regression model. Classical joint-point estimation was performed using an iterative reweighted least squares (IRLS) algorithm, implemented via the segmented function^31^. Initial linear regression models were fitted using the ordinary least squares (OLS) method, and joint-point estimates were refined using the Davies test for significance.^32,33^ Model convergence was assessed through visual inspection of residual plots and diagnostic criteria, including Akaike Information Criterion (AIC) for model selection. Model assumptions were validated by checking for normality of residuals (Shapiro-Wilk test), homoscedasticity (Breusch-Pagan test), and absence of autocorrelation (Durbin-Watson test).

The classical modeling approach does not provide direct uncertainty quantification for the estimated change points. To address this limitation, we implemented a Bayesian joint-point model using the mcp package in R.^34^ The Bayesian approach allows for posterior distribution estimation of all parameters, including the change points, enabling probabilistic interpretation and credible intervals for the breakpoints. The Bayesian model parameters are estimated via Markov Chain Monte Carlo (MCMC) sampling. Prior distributions were assigned to model parameters using a Conjugate-Empirical Bayes approach,^35^ including normal priors for regression coefficients and a uniform prior for joint-point locations. Hyperparameters for the prior distributions were estimated from the classical IRLS estimates.

Model inference was conducted using Hamiltonian Monte Carlo (HMC). Convergence was evaluated through multiple diagnostics, including the Gelman-Rubin diagnostic, effective sample size (ESS), the Rhat statistic, and visual inspection of trace plots. Three independent HMC chains were executed in parallel to assess sampling stability and consistency across chains.

A detailed description of both the mixed effects and joint-point regression models is presented in the supplemental material. In addition, the Bayesian output tables with estimates and 95% Highest Posterior Density Interval (HPDI) or credible intervals are provided in the supplemental material. All analyses were performed using R: A language and environment for statistical computing.^36^ using the lm, segmented, and mcp functions.

### Standard Protocol Approvals, Registrations and Patient Consents

The study was approved by an ethical standards committee on human subject research at Weill Cornell Medicine (*approval #22-12025448*). In accordance with the Declaration of Helsinki, written informed consent was obtained from all study participants.

## Results

### Patient Cohort

This analysis included 29 patients, with 22 females (75.9%). Patient demographics and imaging details are present in Table 1. Most of the cohort had relapsing remitting MS (82.8%). The median number of PRLs per patient was 3.0 [2.0, 4.0]. Overall, 1458 lesions were included in the analysis, of which 97 (6.67%) were PRLs. Disability, measured by EDSS, remained stable across pre- and post-treatment time points. The median number of pre-treatment scans (5.0 [4.0, 6.0]) was similar to post-treatment scans (4.0 [3.0, 6.0]). Figure 2 outlines the longitudinal study design.

**Table 1.** Demographics and Imaging Characteristics.

|  |  |
| --- | --- |
| Number of patients | 29 |
| Sex, n (%) |  |
| Males | 7 (24.1) |
| Females | 22 (75.9) |
| Disease category, n (%) |  |
| CIS | 2 (6.9) |
| RRMS | 24 (82.8) |
| SPMS | 2 (6.9) |
| PPMS | 1 (3.4) |
| Disease duration (years), mean (SD) | 6.79 (6.31) |
| Age, mean (SD) | 40.5 (10.3) |
| EDSS at baseline, median [IQR] | 1.25 [0.0, 2.0] |
| EDSS at treatment initiation, median [IQR] | 1.50 [0.0, 3.0] |
| EDSS at last follow-up, median [IQR] | 1.25 [0.0, 3.6] |
| Prior DMT, n (%) |  |
| Low efficacy | 9 (31.0) |
| High efficacy | 9 (31.0) |
| None | 11 (37.9) |
| T2FLAIR lesion count per patient, median [IQR] | 43.0 [26.0, 65.0] |
| PRL count per patient, median [IQR] | 3.00 [2.00, 4.00] |
| Pre-treatment follow up time (years), median [IQR] | 4.20 [1.18, 5.62] |
| Post-treatment follow up time (years), median [IQR] | 4.09 [3.44, 5.81] |
| Number of pre-treatment scans, median [IQR] | 5.00 [4.00, 6.00] |
| Number of post-treatment scans, median [IQR] | 4.00 [3.00, 6.00] |
| First post-treatment MRI (months), median [IQR] | 9.84 [2.40, 12.12] |

### Source Separation Treatment Analysis for All Lesions

The results of the QSMp mixed effects model estimating the effect of pre- and post-treatment periods for PRLs and non-PRLs are presented in Fig. 3A. As expected, the mean pre-treatment QSMp for PRLs was 33.9 ppb (95% CI: 31.4, 36.5) and was, on average, 3.27 ppb higher than that of non-PRLs (p = 0.001).^6,37,38^ After treatment the reduction in QSMp for PRLs was 2.23 ppb greater than the reduction observed in non-PRLs (p < 0.001). The conditional R² for the model was 0.80.

**Figure 3:**
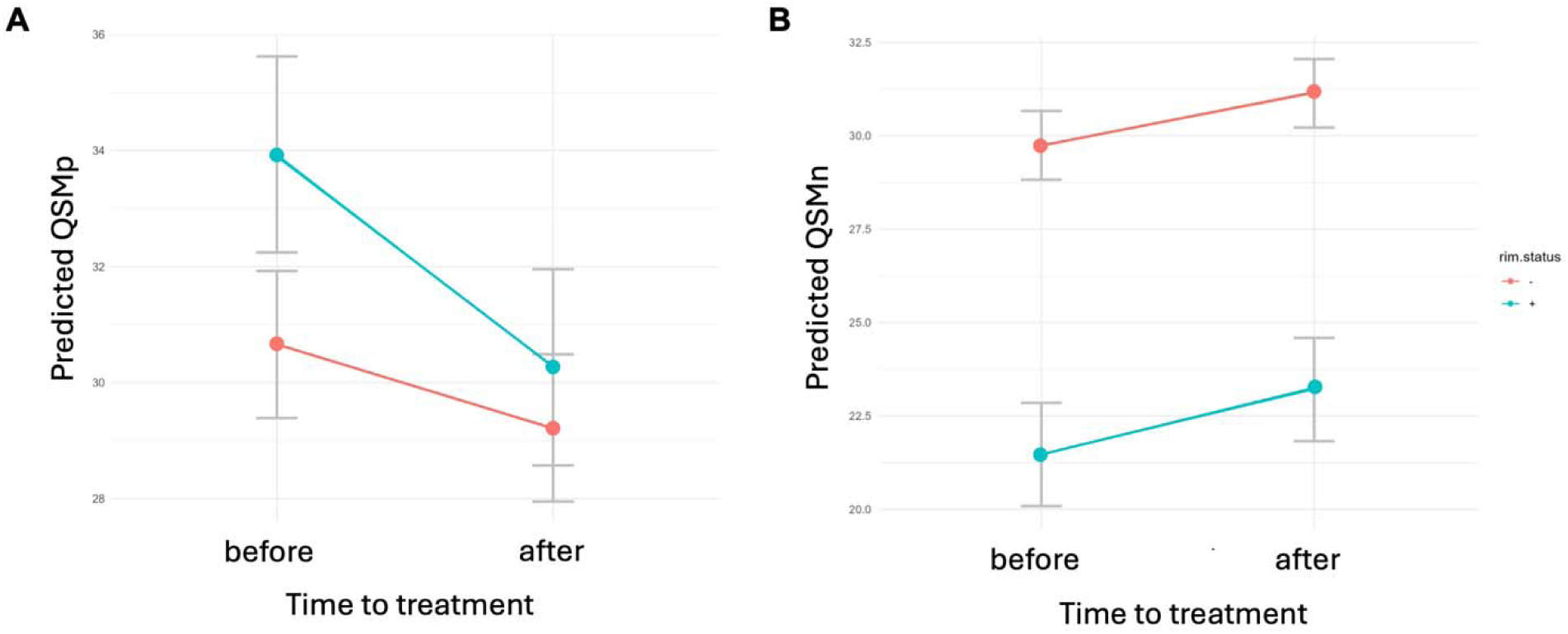
Mixed Effects Model of QSMp and QSMn in PRLs and non-PRLs. 1,361 non-PRLs and 97 PRLs were included both mixed effects models. (A) Mixed effects model of QSMp values in PRLs (blue) and non-PRLs (red) pre- and post-treatment. Pre-treatment QSMp was higher in PRLs (p = 0.001). Post-treatment, PRLs showed an additional 2.23 ppb reduction beyond that in non-PRLs (p < 0.001). (B) Mixed effects model of QSMn values in PRLs (blue) and non-PRLs (red) pre- and post-treatment. Pre-treatment QSMn was lower in PRLs compared to non-PRLs (p < 0.001). Post-treatment changes in QSMn were similar (p = 0.382).

The mixed effects model for QSMn (Fig. 3B) similarly estimated the effect of pre- and post-treatment periods for PRLs and non-PRLs. On average, pre-treatment QSMn for PRLs was 8.27 ppb lower than that of non-PRLs (p < 0.001), consistent with greater demyelination in lesions exhibiting chronic inflammation.^10,39^ The post-treatment increase in QSMn for PRLs was similar to that of non-PRLs (p = 0.382). The conditional R² for the model was 0.741.

### Joint-Point Analysis for PRLs

The Bayesian joint-point regression model suggests that a notable change in the QSMp trajectory for PRLs occurred approximately 4 months after treatment initiation (0.34 years) (Fig. 4A). The 95% HPDI indicates that this change likely took place between treatment initiation and month 7. Prior to the change point, QSMp for PRLs was decreasing at an average rate of –0.44 units per year (95% HPDI: −0.79, −0.09). Following the change, the decline accelerated to –1.08 units per year (95% HPDI: −1.43, −0.80). The model estimates the residual standard deviation (amount of unexplained variability) lies between 8.89 and 9.71 units with 95% probability. This indicates that while the model fits the data well, individual data points tend to deviate from model predictions by roughly 9 units.

**Figure 4:**
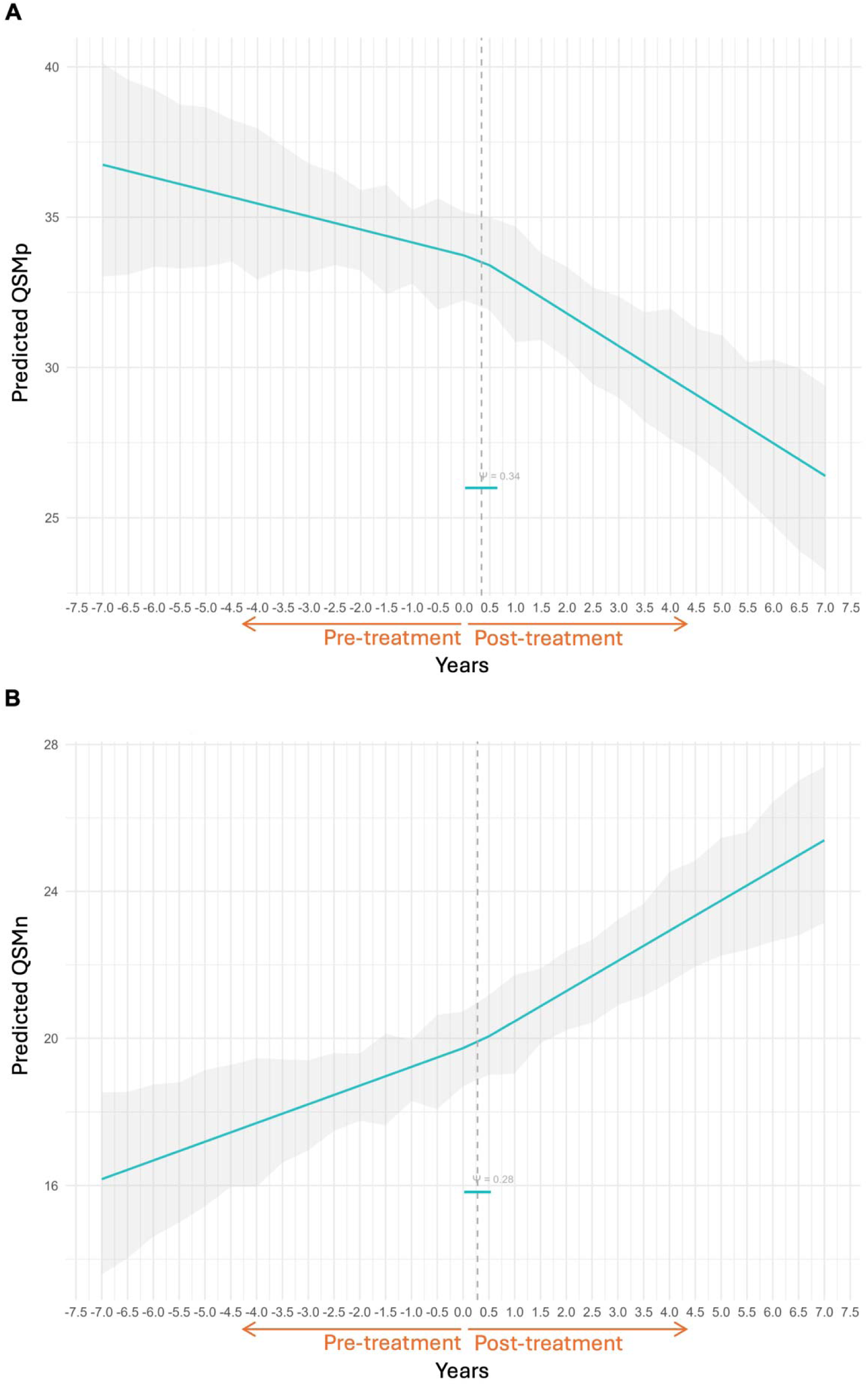
Bayesian Joint-Point Model of QSMp and QSMn trajectories in PRLs. 97 PRLs were included in both Bayesian joint-point models. (A) Longitudinal trajectory of QSMp with estimated change point timing. The shaded grey area shows the 95% confidence interval (not accounting for data correlation). A change point was estimated at 0.34 years (95% credible interval: post-treatment time 0 to month 7). (B) Longitudinal trajectory of QSMn (blue) with estimated change point timing at 0.28 years (95% credible interval: post-treatment time 0 to month 6).

Similarly, the Bayesian joint-point regression model identifies a likely change in the QSMn trajectory for PRLs approximately 3 months after treatment initiation (0.28 years) (Fig. 4B). The 95% credible interval places this shift between treatment initiation and month 6. Before the change, QSMn for PRLs increased at an average rate of 0.51 units per year (95% HPDI: 0.26, 0.76) whereas following the change point, the QSMn increased to a rate of 0.82 units per year (95% HPDI: 0.58, 1.08). The residual standard deviation is estimated to be between 6.09 and 6.66 units with 95% probability, again indicating a reasonably good model fit with modest data variability.

## Discussion

Our study provides the first imaging-based evidence demonstrating a positive impact of Ocrelizumab on PRLs, a key contributor of CNS inflammation in MS. R2*QSM SS of iron revealed a post-treatment reduction in QSMp values, suggesting decreased pro-inflammatory activity within PRLs. Simultaneously, myelin content (QSMn) showed a modest increase, suggesting that while inflammation was attenuated, remyelination may be occurring.

Ocrelizumab is a humanized monoclonal antibody that selectively depletes CD20+ B cells, which play a central role in the pathogenesis of MS. The CD20 glycoprotein is expressed on pre-B and mature B cells but is absent on plasma cells, allowing Ocrelizumab to target pathogenic B cells while preserving antibody-producing B cells.^40^ Upon binding to CD20, Ocrelizumab induces B cells depletion through antibody-dependent cellular cytotoxicity, complement-dependent cytotoxicity, and apoptosis, leading to a profound reduction in circulating and lymphoid-resident B cells.^40^ Notably, Ocrelizumab does not cross the BBB under normal conditions; however, its depletion of peripheral B cells may result in downstream effects on CNS inflammation, including the modulation of microglial and macrophage activity. Several studies have shown that B cell depletion may reduce antigen presentation, pro-inflammatory cytokine release (TNF-α, IL-6, GM-CSF), and CNS immune cell activation.^41^ Since microglia are the dominant CNS-resident antigen-presenting cells in MS, their activation state is tightly linked to interactions with B cells, and the reduced B cell concentrations may consequently lead to decreased activation of brain-derived innate immune cells.^42^

TSPO-PET imaging studies in animal models have demonstrated that Ocrelizumab is associated with reduced microglial activation in both white matter lesions and normal appearing white matter, suggesting a dampening of innate immune activity in the CNS.^43^ Additionally, histopathological analyses have identified ectopic B cell follicles in the meninges, which sustain chronic inflammation and drive neurodegeneration.^44^ These findings highlight the need for further investigation into the downstream effects of Ocrelizumab on CNS inflammation. While Ocrelizumab shows promise in reducing microglial activation, its full impact on chronic lesion pathology remains unclear. Advanced imaging techniques can offer critical in vivo insights into how B cell depletion affects chronic inflammation and lesion progression in MS.

In this study, we used PRLs as a marker of chronic inflammation, as these lesions are known to harbor ongoing inflammatory activity at their edges.^45^ QSM, which leverages the differential magnetic properties of tissue, can accurately detect PRLs^6,37,38^ and quantify composite paramagnetic iron deposition, such as ferritin or hemosiderin, as well as diamagnetic water content within myelin.^16^ However, to more effectively isolate the contributions of iron and myelin, we applied the R2*QSM SS algorithm, developed by our group, to disentangle these signals. This approach utilizes GRE magnitude decay modeling and phase-based reconstruction, allowing for more precise detection of iron (QSMp) and myelin loss/recovery (QSMn).^19^ This validated technique^18^ is particularly valuable for assessing PRLs, given the increased demyelination in PRLs^39^ may obscure the effects of iron on susceptibility values.

In contrast to our findings, prior QSM-based studies evaluating Ocrelizumab’s effect on PRLs did not detect a significant treatment response.^46^ This may reflect inherent limitations of QSM, which measures a combined signal from iron and myelin, potentially masking subtle lesion changes. Additionally, short follow-up durations may fail to capture long-term lesion dynamics. To address these issues, our study leveraged all available imaging data from both pre- and post-treatment periods, as using each lesion as its own control requires extended observation to detect meaningful shifts in its trajectory. In the current study, we found that QSM demonstrated a shift after treatment (data not shown), suggesting that a longer observation may be required. We further used QSMp rather than conventional QSM to assess post-treatment inflammation, which may provide greater sensitivity. While the precise timing of the observed shift is difficult to determine due to variability in MRI scheduling (median: 9.8 months post-treatment), our findings consistently indicate that the change occurs following treatment initiation.

Although our study provides novel insights into the effects of Ocrelizumab on chronic CNS inflammation in MS by leveraging R2*QSM SS, several limitations must be acknowledged. First, the reliability of current susceptibility SS methods,^47^ including our R2*QSM algorithm, for myelin quantification may be influenced by the fiber orientation. The myelin sheath consists of concentric lipid bilayers tightly wrapped around cylindrical axons, which are bundled together, leading to orientation-dependent magnetic susceptibility.^48–50^ Therefore, caution is necessary to avoid overinterpretation of QSMn results. New susceptibility SS approaches are in development that account for these effects through detailed microscale signal modeling.^51^ Also, the small cohort size limited our ability to control for patient demographics or clinical variables, such as disease category or previous disease-modifying treatments, as well as our ability to assess treatment effects on disability. Moreover, because disability scores remained stable over time, we could not evaluate a potential link between reduced PRL inflammation and changes in disability, even in an exploratory analysis. Larger studies are needed to determine whether dampening PRL inflammatory activity leads to measurable clinical improvement. Second, we lacked information about lesion age, meaning some PRLs may have already been in later stages of their natural decline.^52^

We observed that the joint-points for QSMp and QSMn occurred at approximately the same time. Logically, remyelination might not be expected to occur at the same time as inflammation reduction; typically, a delay in remyelination would be anticipated. While some coupling between QSMp and QSMn is expected due to the modeling, three points warrant emphasis: 1) as mentioned, the exact timing of the trajectory shift cannot be determined, but it clearly occurs after treatment initiation; 2) QSMn should be interpreted cautiously as a biomarker for myelin; and (3) a similar increase in QSMn was observed in both PRLs and non-PRLs, in contrast to significantly different changes seen with QSMp. Thus, the divergent pattern observed in the mixed effects model suggests some uncoupling of these measures. Moreover, our QSMn results indicate that if remyelination is occurring, it may be affecting all chronic lesions. This finding is consistent with previous myelin water imaging studies in clinical trial patients treated with Ocrelizumab, supporting the possibility that Ocrelizumab promotes remyelination.^53^

In conclusion, this study provides the first evidence that Ocrelizumab significantly reduces iron content within PRLs, indicating a decline in pro-inflammatory activity. The modest and non-significant increase in myelin content suggests that remyelination may be a slower process and occurs broadly among all lesions. The treatment-related change in lesion susceptibility further highlights the potential of PRLs as imaging biomarkers for monitoring therapeutic response in MS, warranting further longitudinal studies to validate their clinical utility in guiding treatment decisions. Future research should focus on expanding the application of susceptibility SS algorithms to larger patient cohorts to better characterize the impact of current DMTs on chronic inflammation within PRLs. Investigating the mechanistic link between B cell depletion and CNS microglial activity could provide deeper insight into how adaptive immune modulation influences innate immune-driven neurodegeneration. These efforts will be critical in guiding therapeutic strategies aimed at mitigating disease progression in MS.

## Data Availability

Raw data were generated at Weill Cornell Medicine. Derived data supporting the findings of this study are available from the corresponding author on request.

## Funding

This study was funded by Genentech.

## Competing Interests

The authors report no conflicting interests.

## Notes

### Competing Interest Statement

The authors have declared no competing interest.

### Funding Statement

This study was funded by Genentech (ML44547)

### Author Declarations

IRB of Weill Cornell Medicine gave ethical approval for this (IRB #22-12025448)

