## Supplemental Materials for "Quantifying the Impact of Ocrelizumab on Paramagnetic Rim Lesions in Multiple Sclerosis"

**Supplemental Material**

We used a model-based approach to study the trajectory of chronic inflammation in PRLs. We conducted a longitudinal analysis of changes in susceptibility across both iron and myelin components, utilizing the R2*QSM source separation framework. The model-based approach includes a mixed effects model and a joint-point regression model.

**A. Mixed Effects Model**

We used mixed effects models [1] to estimate the effect of pre- and post-treatment periods on QSM biomarkers while accounting for multiple lesions per patient. The model was fitted using restricted maximum likelihood (REML), and its fit was assessed through likelihood ratio tests, AIC, and residual diagnostics. Random intercept variance estimates were examined to ensure appropriate within-subject correlation modeling. We estimated the following questions:

${QSMp}_{ij}=\beta_{1} {TRT}_{ij}+ \beta_{2} {PRL}_{ij}+ + \beta_{12} {TRT}_{ij}* {PRL}_{ij}+b_{oj}+b_{oij}+\epsilon_{ij}$	(1)

${QSMn}_{ij}=\beta_{1} {TRT}_{ij}+ \beta_{2} {PRL}_{ij}+ + \beta_{12} {TRT}_{ij}* {PRL}_{ij}+b_{oj}+b_{oij}+\epsilon_{ij}$	(2)

Where:

- ${QSMp}_{ij}$is the QSMp susceptibility (response) for lesion i patient j. Similarly, ${QSMn}_{ij}$is the QSMn susceptibility for lesion i patient j
- $\beta_{1}$ is the fixed effect of pre_post_treatment effect (${TRT}_{ij}$) for lesion i, patient j.
- $\beta_{2}$ is the fixed effect of lesion type (${PRL}_{ij}=1$ if lesion i, patient j is a PRL lesion versus 0 otherwise)
- $\beta_{12}$ is the fixed effect of the interaction between pre_post_treatment status (${TRT}_{ij}$) and lesion type (${PRL}_{ij}$)

- $b_{oj} \sim N(0, \sigma_{subj}^{2})$ is the random intercept for patient j
- $b_{oij} \sim N(0, \sigma_{lesion}^{2})$ is the random intercept for lesion i within patient j
- $\epsilon_{ij} \sim N(0, \sigma^{2}$) is the residual error

Mixed-effects models offer several key advantages, especially when analyzing data with complex structures. One major benefit is their ability to account for both fixed effects (pre_post treatment and lesion type) and random effects (multiple lesions per patient). By modeling random variation appropriately, mixed-effects models improve the accuracy of estimates and reduce the risk of inflated Type I error rates. They also handle our unbalanced (different number of lesions per patient) data well without requiring listwise deletion. Overall, mixed-effects models provide a flexible and powerful framework for analyzing QSM data at the lesion level.

**B. Mixed Effects Model Estimates**

**QSMp Model**

| **Parameter** | **Estimate** | **95% C.I** | **p-value** |
| --- | --- | --- | --- |
| pre_treatment effect | 30.66 | (28.76, 32.55) | <0.001 |
| post_treatment effect | 29.22 | (27.32, 31.11) | <0.001 |
| PRLs | 3.27 | (1.36, 5.18) | <0.001 |
| post_treatment effect x PRLs | -2.23 | (-2.98 , -1.48) | <0.001 |

**Table 1.** Fixed effects estimates from the QSMp linear mixed-effects model. Reported values include the estimated coefficients (Estimate), 95% confidence intervals (95% C>I), and associated p-values (p). Confidence intervals reflect the uncertainty in the parameter estimates. Statistical significance was assessed at the 0.05 level.

**QSMn Model**

| **Parameter** | **Estimate** | **95% C.I** | **p-value** |
| --- | --- | --- | --- |
| pre_treatment effect | 29.74 | (28.37, 31.11) | <0.001 |
| post_treatment effect | 31.13 | (29.76, 32.51) | <0.001 |
| PRLs | -8.27 | (-10.03, -6.52) | <0.001 |
| post_treatment effect x PRLs | 0.34 | (-0.43,  1.12) | 0.382 |

**Table 2**. Fixed effects estimates from the QSMn linear mixed-effects model. Reported values include the estimated coefficients (Estimate), 95% confidence intervals (95% C>I), and associated p-values (p). Confidence intervals reflect the uncertainty in the parameter estimates. Statistical significance was assessed at the 0.05 level.

Diagnostic checks of the linear mixed-effects models revealed that the residuals were approximately normally distributed, as evidenced by Q-Q plots, and showed no systematic patterns when plotted against fitted values, supporting the assumption of homoscedasticity. No influential outliers were detected based on standardized residuals. Both model's explanatory power was assessed using the conditional $R^{2}$ , which accounts for both fixed and random effects. The conditional $R^{2}$ for the QSMp model was 0.80, indicating that the model explains 80% of the variance in the outcome after accounting for both patient-level and lesion-level variation. Similarly, the conditional $R^{2}$ for the QSMn model was 0.74. Overall, all residual diagnostics and fit indices suggest that both models appropriately capture the structure of the data.

**C. Joint-point regression models**

A linear joint-point regression model was used to estimate the change points and slopes of the QSM trajectories for PRLs over time (see Equations 3 and 4). This joint-point model enables the detection of structural changes in the time variable ($t_{ij}$), identifying breakpoints where the trajectory undergoes a shift [2]. In this model, time is defined in years (or fractions of a year), with negative values representing the period before treatment and positive values indicating the period after treatment.

${QSMp}_{ij}\left( t_{ij} \right)=\beta_{0}+b_{oj}+\beta_{1} t_{ij}+\beta_{2} {( t_{ij}- \tau)}_{+}+\epsilon_{ij}$	(3)

${QSMn}_{ij}\left( t_{ij} \right)=\beta_{0}+b_{oj}+\beta_{1} t_{ij}+\beta_{2} {( t_{ij}- \tau)}_{+}+\epsilon_{ij}$	(4)

${QSMp}_{ij}$is the QSMp susceptibility (response) for lesion i patient j at time t. Similarly, ${QSMn}_{ij}$is the QSMn susceptibility for lesion i in patient j, $\beta_{0}$ is the fixed intercept, $b_{oj} \sim N(0, \sigma_{subj}^{2})$ is the random intercept for patient j, $\beta_{1}$is the slope before the change point, $\beta_{2}$ is the change in slope after the change point, τ represents the change point, ${( t_{ij}- \tau)}_{+}= max\{0, t_{ij}- \tau\}$ represents the “hinge” or segmented term, active only after the change point, and $\epsilon_{ij} \sim N(0, \sigma^{2}$) is the residual error.

We implemented both a classical and Bayesian joint-point regression model. The classical implementation estimated change-points using an iterative reweighted least squares algorithm (IRLS), as implemented in the segmented function [3]. Initial linear regression models were fitted using the ordinary least squares (OLS) method, and change-point estimates were refined using the Davies test for significance [4,5]. Model convergence was assessed through visual inspection of residual plots and diagnostic criteria, including Akaike Information Criterion (AIC) for model selection. Model assumptions were validated by checking for normality of residuals (Shapiro-Wilk test), homoscedasticity (Breusch-Pagan test), and absence of autocorrelation (Durbin-Watson test).

A disadvantage of the classical approach is that it does not provide direct uncertainty quantification for the estimated change points. To address this limitation, we implemented a Bayesian change-point model using the mcp package in R [6]. The Bayesian approach allows for posterior distribution estimation of all parameters, including the change points, enabling probabilistic interpretation and credible intervals for the breakpoints. The Bayesian model parameters are estimated via Markov Chain Monte Carlo (MCMC) sampling. Prior distributions were assigned to model parameters using a Conjugate-Empirical Bayes approach [7], including normal priors for regression coefficients and a uniform prior for change-point locations. Hyperparameters for the prior distributions were estimated from the classical IRLS estimates. Model inference was conducted using Hamiltonian Monte Carlo (HMC). Convergence was evaluated through multiple diagnostics, including the Gelman-Rubin diagnostic, effective sample size (ESS), the Rhat statistic, and visual inspection of trace plots. Three independent HMC chains were executed in parallel to assess sampling stability and consistency across chains.

All analysis was performed using R: A language and environment for statistical computing [8] using the lm(), segmented(), and mcp() functions.

**D. Bayesan Joint-point regression model Estimates**

**QSMp- Posterior Estimates and 95% HPDI**

| **Parameter** | **Posterior Mean** | **HPDI Lower** | **HPDI Upper** | **Rhat** | **EB Prior**  **Mean** | **EB Prior**  **SD** |
| --- | --- | --- | --- | --- | --- | --- |
| τ | 0.3427 | 0.0223 | 0.6444 | 1.0001 | 0.25 | 0.25 |
| $\beta_{0}$ | 33.7184 | 32.8578 | 34.6204 | 1.0008 | 33 | 12 |
| β₁ | -0.4352 | -0.7925 | -0.0866 | 1.0003 | -0.46 | 1 |
| β₂ | -1.077 | -1.4315 | -0.6999 | 1.0006 | -1.04 | 1 |
| σ (Residual ) | 9.3145 | 8.8943 | 9.7124 | 0.9999 | 25 | 10 |

**Table 3.** Bayesian posterior estimates for model parameters corresponding to QSMp. The table reports the posterior mean estimates, the 95% Highest Posterior Density Interval (HPDI), the Rhat statistics as convergence diagnostic measurement, and prior information from the empirical Bayes (EB) estimates based on the classical model. The prior means were derived from estimates obtained through a classical model, while the prior standard deviations (SD) were inflated to ensure a flatter prior distribution.

**QSMn- Posterior Estimates and 95% HPDI**

| Parameter | Posterior Mean | HPDI Lower | HPDI Upper | Rhat | EB Prior  Mean | EB Prior  SD |
| --- | --- | --- | --- | --- | --- | --- |
| τ | 0.2793 | 0.0217 | 0.5395 | 1.002 | 0.20 | 0.20 |
| $\beta_{0}$ | 19.7330 | 19.0937 | 20.31280 | 1.001 | 14.88 | 10 |
| β₁ | 0.5084 | 0.2564 | 0.7558 | 1.001 | 0.45 | 1 |
| β₂ | 0.8211 | 0.5763 | 1.0843 | 1.000 | 0.81 | 1 |
| σ (Residual ) | 6.3812 | 6.0870 | 6.660 | 1.001 | 20 | 10 |

**Table 4.** Bayesian posterior estimates for model parameters corresponding to QSMn. The table reports the posterior mean estimates, the 95% Highest Posterior Density Interval (HPDI), the Rhat statistics as convergence diagnostic measurement, and prior information from the empirical Bayes (EB) estimates based on the classical model. The prior means were derived from estimates obtained through a classical model, while the prior standard deviations (SD) were inflated to ensure a flatter prior distribution.

Posterior estimates represent the central tendency of the model parameters after accounting for prior distributions, while the HPDI provides the range within which the true parameter value lies with 95% probability. The Rhat statistic is used to assess convergence in Bayesian models by comparing the variance within chains to the variance between chains. An Rhat value close to 1 indicates that the model has converged, suggesting that the chains are mixing well and exploring the parameter space effectively. Prior information reflects the EB estimates, which were derived from classical IRLS fitting before the Bayesian updating process.

**E. Bayesian Hypothesis Testing**

Hypotheses about parameter values can be tested using the p and Bayes Factors (BF) calculated from the Savage-Dickey density ratio [9]. A value of 𝑝 close to 1 indicates strong evidence in favor of the specified hypothesis. A Bayes factor (BF) greater than 1 strengthens belief in the point hypothesis by a factor of approximately ‘BF’ relative to prior beliefs. Conversely, the inverse (i.e., 1 − p and 1/BF) is the evidence in favor of the alternative.

**QSMp- Hypothesis**

| **Null Hypothesis** | **p** | **BF** |
| --- | --- | --- |
| τ >0 | 1.0000 | Inf |
| -0.5 <β₁ <0 | 0.6406 | 1.7829 |
| β₂ <2* β₁ | 0.6744 | 2.0716 |

**Table 5.** All 𝑝 are greater than 0.6 indicating a strong evidence in favor of the specified hypothesis. All BF>1 indicates a strong belief in the point hypothesis by a factor of approximately ‘BF’ relative to prior beliefs.

**QSMn- Hypothesis**

| **Null Hypothesis** | **p** | **BF** |
| --- | --- | --- |
| τ >0 | 0.9701 | 33.3511 |
| 0< β₁ < 0.75 | 0.5846 | 1.4077 |
| β₂ > 1.5* β₁ | 0.9855 | 68.2307 |

**Table 6.** All 𝑝 are greater than 0.58 indicating a strong evidence in favor of the specified hypothesis. The fact that all BF>1, strengthens belief in the point hypothesis by a factor of approximately ‘BF’ relative to prior beliefs

**F. Bayesian Convergence Diagnosis**

Model inference was conducted using Hamiltonian Monte Carlo (HMC). Convergence was evaluated through multiple diagnostics, including the Gelman-Rubin, effective sample size (ESS), the Rhat statistic, and visual inspection of trace plots. Three independent HMC chains were executed in parallel to assess sampling stability and consistency across chains.

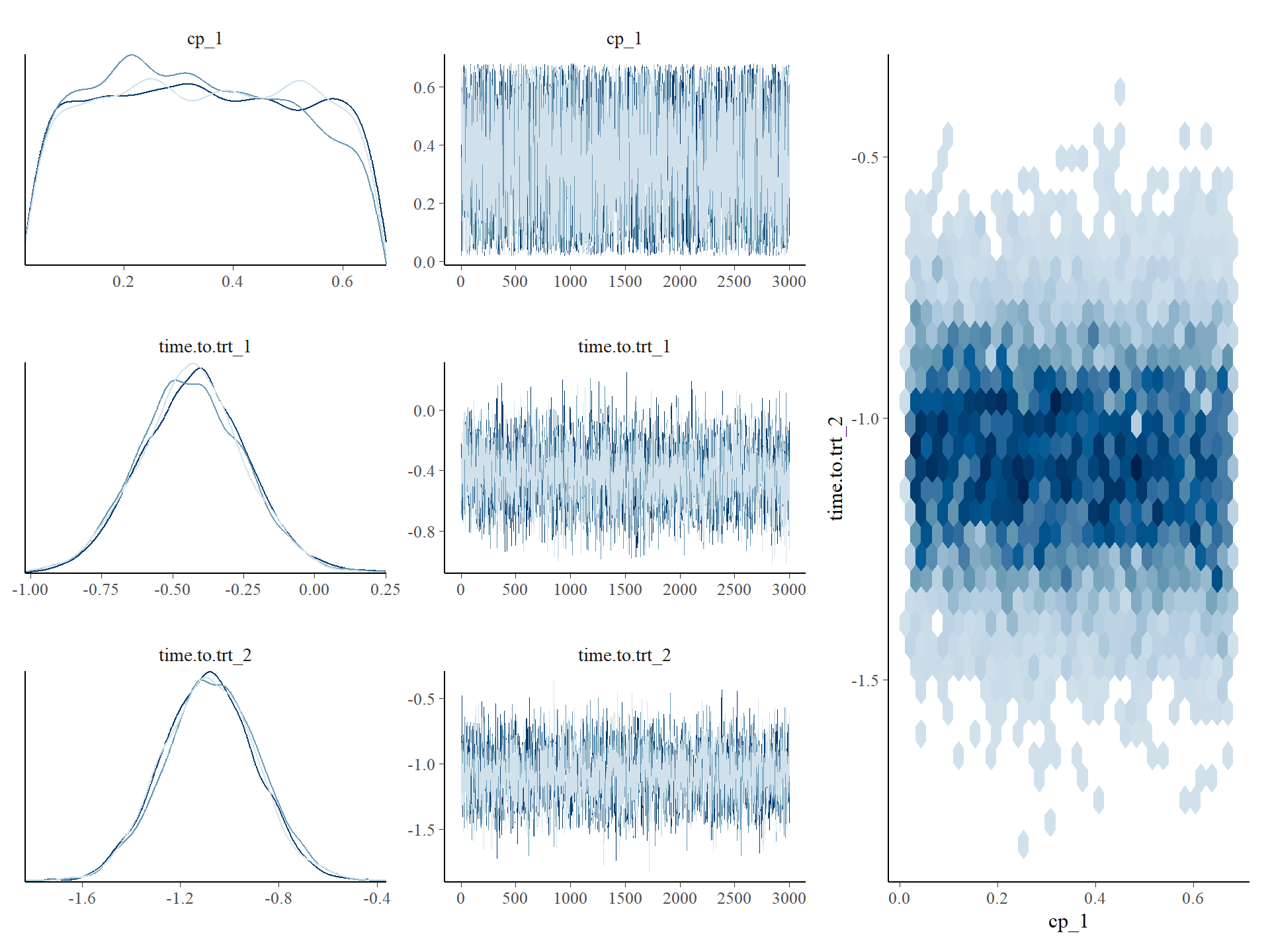

**Figure 1.** QSMp Convergence Diagnosis. Left: Marginal posterior distributions and trace plots for a subset of model parameters (organized by row), with three MCMC chains distinguished by color. Diagnostics indicate satisfactory mixing and convergence across chains, suggesting stable posterior sampling. Right: Joint posterior density plot illustrating the bivariate relationship between the estimated change point and the slope parameter of the subsequent segment, highlighting potential posterior dependency or trade-offs between these estimates. cp_1 represents the change points, time.to.trt1 represents the first slope and time.totr2 indicates the slope after change point.

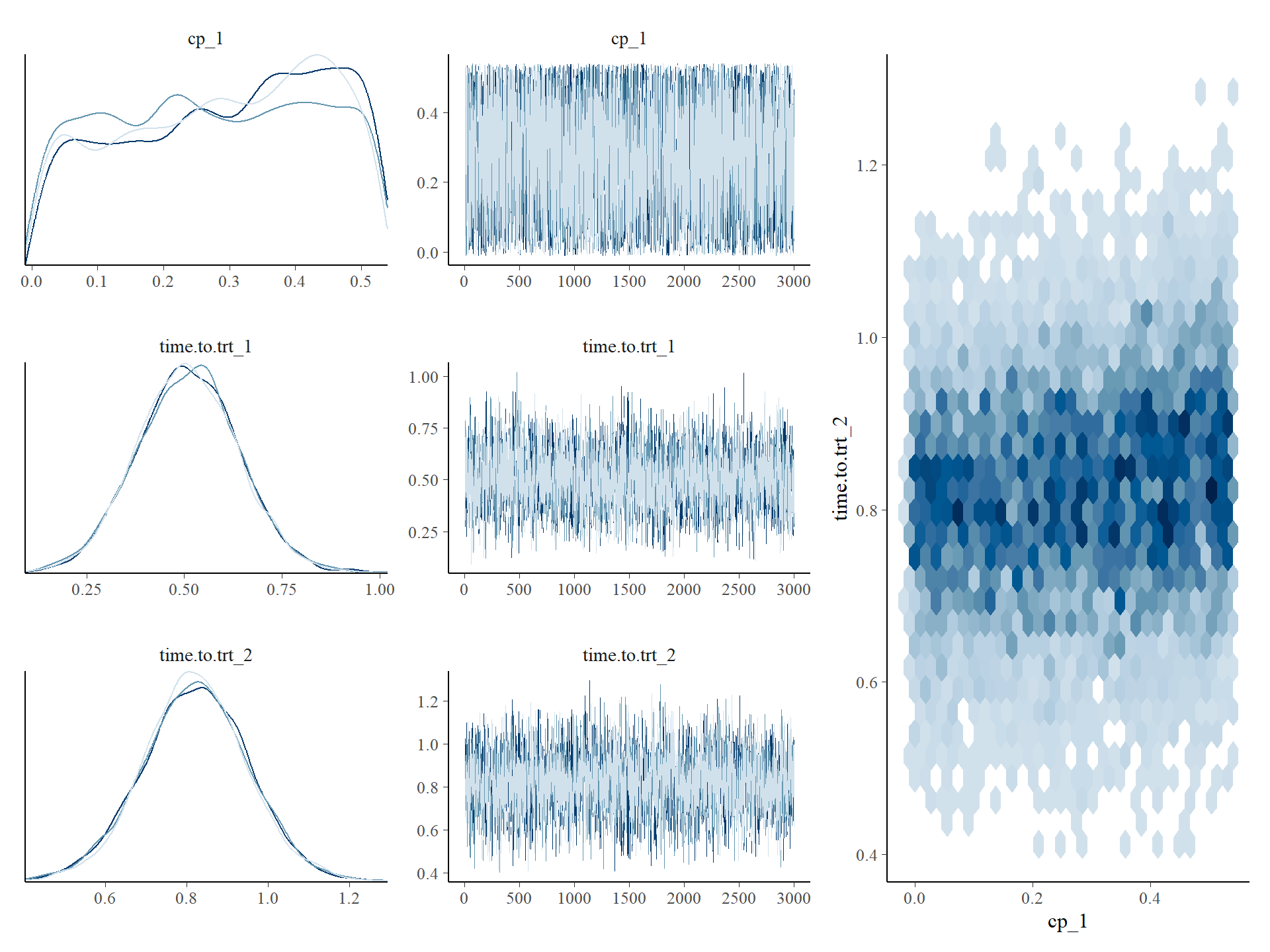

**Figure 2.** QSMn Convergence Diagnosis. Left: Marginal posterior distributions and trace plots for a subset of model parameters (organized by row), with three MCMC chains distinguished by color. Diagnostics indicate satisfactory mixing and convergence across chains, suggesting stable posterior sampling. Right: Joint posterior density plot illustrating the bivariate relationship between the estimated change point and the slope parameter of the subsequent segment, highlighting potential posterior dependency or trade-offs between these estimates. cp_1 represents the change points, time.to.trt1 represents the first slope and time.totr2 indicates the slope after change point.
